# Research output is low, but increasing, and focuses on public health at new universities in southern Ethiopia

**DOI:** 10.1101/2021.10.01.21264396

**Authors:** Bernt Lindtjørn, Fekadu Massebo, Taye Gari, Solomon Hailemariam Tesfaye

**Author notes:** Correspondence to: Bernt Lindtjørn. E-mail addresses: Bernt Lindtjørn Fekadu Massebo; Taye Gari Solomon Hailemariam.

## Abstract

**Objective:** The aim of this work is to describe the developments in research publication at three new universities in southern Ethiopia.

**Design:** Bibliometric analysis.

**Methods:** We retrieved references from 1998 to July 2021 from the Web of Science database using the authors’ affiliation. English language publications and peer-reviewed journals were included. The bibliometric analyses were conducted by using Web of Science and the bibliometrix package (version 3.1) under R version 4.0.5.

**Results:** We reviewed 1019 scientific papers, and there was a substantial increase in the number of publications over the years, especially since 2016. The publications were in 304 different journals with 2606 authors; the number of documents per author was 0·39. Most of the studies were descriptive, 17 (1·7%) were clinical trials, 60 (5·9%) were case-control studies, and 83 (8·1%) were cohort studies. Topics related to public health were the most often studied. The average citations per publication was 9·1. The most frequently cited papers occurred with international collaboration. A total of 886 (84%) publications were “All Open Access” and only 5% of the papers were published in Ethiopian journals. We identified ten groups that maintained scientific production for 8 years or more, mainly in research on malaria and vector borne diseases, nutrition, microbiology, and various public health issues. One of seven papers were published with support from external funding, and with collaborative links with the United States, Europe, and New Zealand.

**Conclusions:** There has been a remarkable increase in health research at the three universities. The institutions should enhance the research culture, strengthening their health research capacity by encouraging good research practice and ensuring connections between health research and implementation.

## Introduction

More than 60 years since most universities in sub-Saharan Africa were founded, a recent review has underlined the modest record of African universities.^1^ Whereas countries in sub-Saharan Africa account for 14% of the global population, they contribute less than 1% of the research expenditure.^1^ Although the research output may have increased in recent years, in 2007, Ethiopia spent only 0·2% of its gross national product on research.^2^ A recent analysis of research collaboration between European and African universities showed that the largest research collaboration is between Europe and South Africa, probably because of the location of research institutions that are both academically good and have adequate research infrastructure.^3^ In line with these findings, most of the publications are from South Africa and Northern Africa.^4^ Countries such as Nigeria and Kenya are among the sub-Saharan African countries that produced the most research.^5^

According to the uniRank database, there were 1225 officially recognized higher-education institutions in Africa in 2020.^6^ Less than half of them are public institutions. Ethiopia now has more than 50 universities, with most of them being less than 20 years old. We analysed publications from three of the younger universities in southern Ethiopia and believe that they are typical of evolving and growing institutions in low-income countries.

Systematic bibliometric analyses can be useful analytical tools to gain a better understanding of the research output of an institution, as well as the research pattern and collaboration at these institutions. However, very few systematic assessments have been done regarding scientific literature production in Ethiopia,^7 8^ and to our knowledge, none include southern Ethiopia. Nevertheless, there have been several bibliographies on specific health problems that included Ethiopia,^9^ as well as topics such as HIV/AIDS and tuberculosis.^10^ During the last 15 to 20 years, newer universities have increasingly collaborated with research institutions in the United States and Europe. It could be worthwhile to evaluate if these collaborations have brought about lasting changes at these universities, causing them to become more independent and to increase their research.^11 12^

Such institutions are expected to provide education and to conduct research that can improve the health of the populations living in their area. The aim of this work is to describe the developments in research publication at universities in Hawassa, Arba Minch, and in Dilla during the past 15 years. Our focus is mainly on the scientific production and networks originating from these institutions.

## Methods

### Background of universities

The universities in Hawassa (uniRank rank 3870 in the world in 2020), Arba Minch (rank 4237), and Dilla (Rank 20086) in southern Ethiopia are young, and they have been functioning as universities for 17–23 years.^6^

Hawassa University was established in 1998 and is the oldest of the universities in southern Ethiopia. It has a well-developed infrastructure and administration, and it has some experience in collaborating with universities outside Ethiopia. In 2021, the College of Health Sciences had 19 PhD holders and 169 staff with master’s degrees engaged in teaching and research activities. In the same year, there were 1085 undergraduate students (medical and paramedical), 496 master students, 161 resident doctors, and 24 PhD students.

Arba Minch University was established in 2004. Health-related research is carried out at the College of Health Sciences and College of Biology. Health related research is mainly carried out at the College of Medicine and Health Sciences, and College of Natural Sciences in particular in the Department of Biological Sciences. Of the total 271 academic staffs, 36 are PhD holders and 140 are MSc and 7 are Bachelor degree.

Dilla University (DU) was established in 2006. Of the 293 staff members in the College of Health Sciences in July 2021, six are PhD holders, 128 have a master’s degree, 122 have a bachelor’s degree, and 37 are medical doctors. Currently, there are 943 students. From these, 106 are master’s students. The college does not have a PhD programme.

Similar data were collected from a small-to medium-sized European university in Bergen, Norway. The University of Bergen (uniRank 245) is a medium-sized, internationally recognised research university with 18 500 students, of whom around 2000 are international students.

### Data collection and analysis

The bibliographic data for this analysis were retrieved from the Web of Science database. The references were extracted from the Web of Science collection on July 20, 2021, primarily using the authors’ affiliation. English language publications and peer-reviewed journals were included. The timespan was for the lifespan of the institutions: Hawassa University (1998–2021), Arba Minch University (2004–2021), and Dilla University (2006– 2021).

Altogether, these three institutions published 1064 publications in peer-reviewed journals, with 41 of the papers being part of large international collaborations on estimating different burdens of global disease. Our primary aim was to analyse the performance and institutional collaborations of these three universities over their lifespan, and 1019 publications were included in our study.

The bibliometric analyses were conducted by using the available tools in Web of Science and the bibliometrix package (version 3.1) under R version 4.0.5, using the Biblioshine software for visualization.^13^

## Results

A total of 1019 scientific papers were reviewed in our analysis, which included 928 research articles and 91 reviews. A substantial increase in the number of publications occurred over the years (Figure 1), especially since 2016. The rate of increase was similar at all three institutions. There were 606 articles from Hawassa University, 240 from Arba Minch University, and 207 from Dilla University. Twenty-eight of the papers were from both Hawassa University and DU, 23 from Hawassa University and Arba Minch University, and 9 from Arba Minch University and Dilla University.

**Figure 1.**
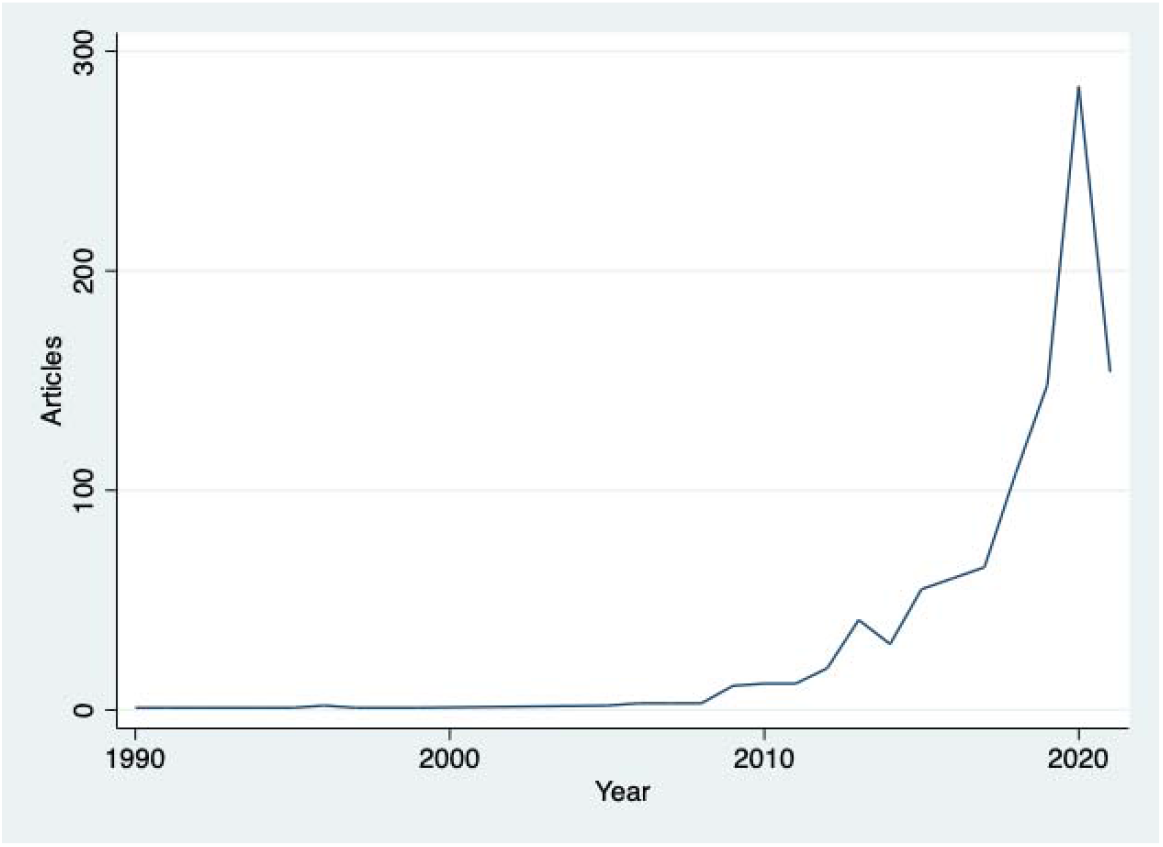
Number of publications per year at the universities in Hawassa, Arba Minch, and Dilla.

The publications were in 304 different journals with 2606 authors (average 2.6 authors per paper), and twenty-four (2·3%) of the papers were written by a single author. The number of documents per author was 0·39.

Most of the studies were descriptive surveys. There were 17 (1·7%) clinical trials, 60 (5·9%) of the papers were case-control studies, and 83 (8·1%) were cohort studies. The number of meta-analysis papers were 79 (7·7%), with 55 (70%) meta-analyses published since 2020.

The average citations per publication was 9·1. The most frequently cited countries were the United States (78 citations per article), New Zealand (44·7), Germany (17·7), and Norway (14·9). Ethiopia was cited 6·9 times per article.

Topics related to public health were the most often studied, and the ten most common categories (91% of all publications; Web of Science categories) are listed in Table 1.

**Table 1.**
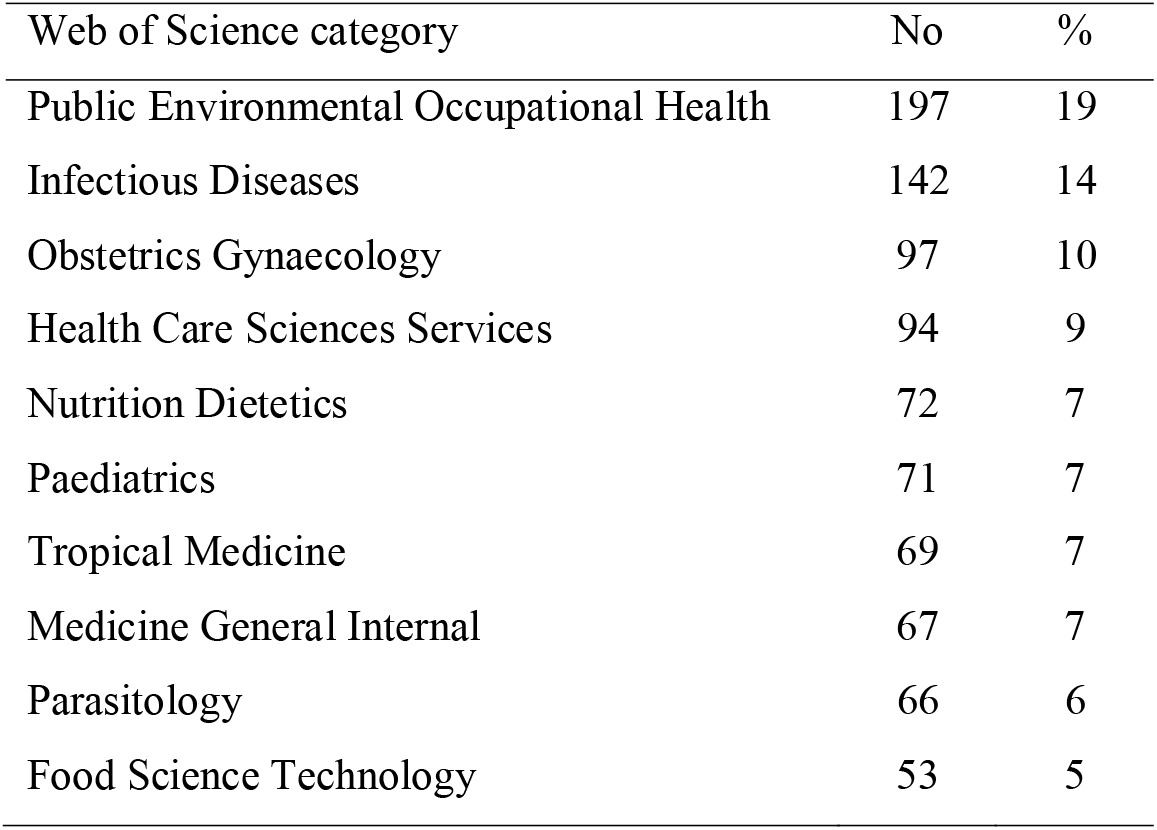
Web of Science categories of publications

| Web of Science category | No | % |
| --- | --- | --- |
| Public Environmental Occupational Health | 197 | 19 |
| Infectious Diseases | 142 | 14 |
| Obstetrics Gynaecology | 97 | 10 |
| Health Care Sciences Services | 94 | 9 |
| Nutrition Dietetics | 72 | 7 |
| Paediatrics | 71 | 7 |
| Tropical Medicine | 69 | 7 |
| Medicine General Internal | 67 | 7 |
| Parasitology | 66 | 6 |
| Food Science Technology | 53 | 5 |

### Sources of publications

A total of 886 (84%) publications were “All Open Access.” The ten most common journals, accounting for 28% of all publications, are listed in Table 2. Only 5% of the papers were published in Ethiopian journals.

**Table 2:** Most used journals

| Sources | Articles | % |
| --- | --- | --- |
| BMC Public Health | 57 | 6 |
| BMC Infectious Diseases | 38 | 4 |
| BMC Pregnancy and Childbirth | 37 | 4 |
| Ethiopian Journal of Health Sciences | 34 | 3 |
| Malaria Journal | 25 | 2 |
| International Journal of Surgery Open | 23 | 2 |
| BMJ Open | 19 | 2 |
| BMC Paediatrics | 18 | 2 |
| HIV Aids-Research and Palliative Care | 18 | 2 |
| Ethiopian Journal of Health Development | 17 | 2 |
| Reproductive Health | 17 | 2 |

In 186 publications (21%), the corresponding author was from a country outside Ethiopia. Collaborators on these publications were most frequently from the United States, Australia, Norway, Belgium, and Canada.

### Research groups

We analysed each of the researcher’s production over time and found that ten groups had maintained scientific production for 8 years or more. They included research groups on malaria and vector borne diseases, nutrition, microbiology, and various public health issues. Two of the groups had a long-standing international collaboration and included several staff from the three universities. Four of these leading researchers left their institutions, with three going to Addis Ababa and one leaving the country.

### International collaboration

Although the Web of Science does not provide much information about the research funding, 141 (14%) articles were published with some support from external funding; 8% from the United States, 2% from the European Union, 2% from Norway, and 1% from others. The remaining publications were funded by the Ethiopian institutions.

The pattern of international collaboration follows the funding, and as seen from the map in Figure 2, the strongest collaborative links were with the United States, the United Kingdom, Norway, Belgium, Sweden, and New Zealand.

**Figure 2.**
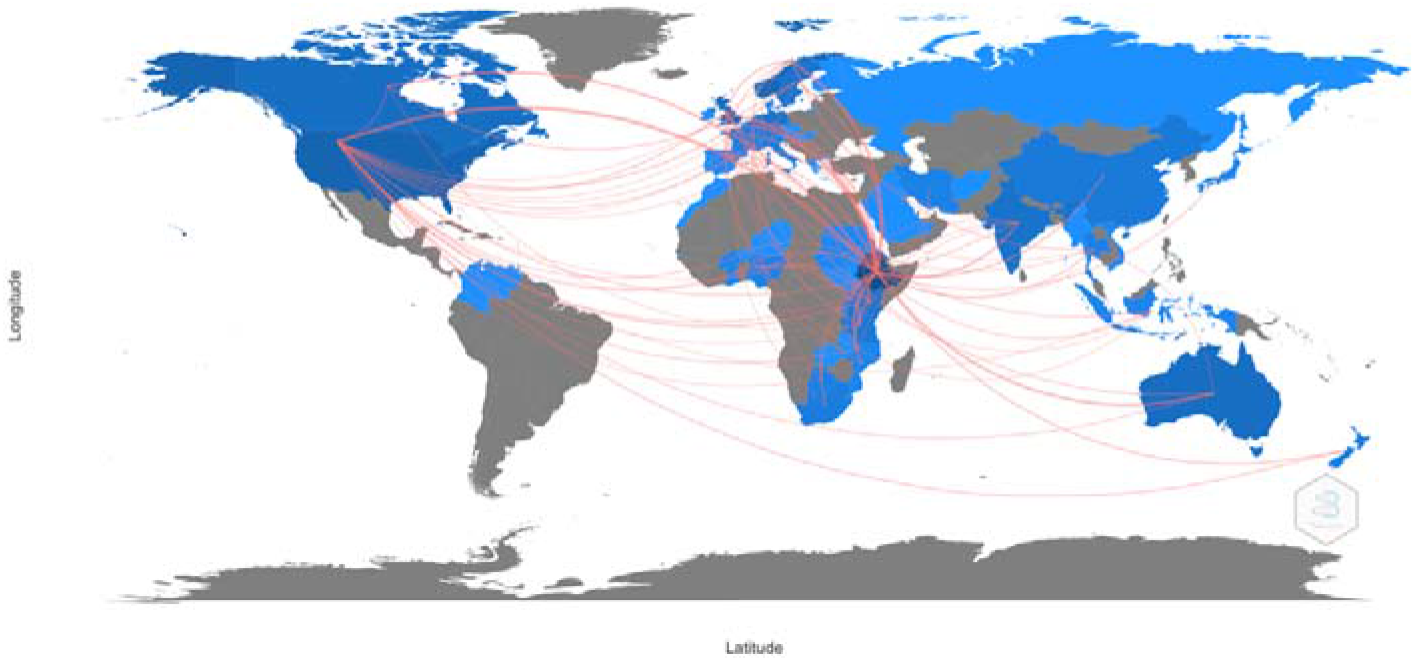
Map of the world showing collaborations of the three universities in southern Ethiopia.

#### Comparison with a medium-sized European university

Similar data collection was done for a medium-sized European university in Bergen, Norway. The selection of articles was done as described for the Ethiopian universities. The research output at the University of Bergen was 17 times higher than that of the three universities from southern Ethiopia combined. The type of research at the University of Bergen also differed from that in the Ethiopian universities, with the Norwegian institution focusing on clinical research and laboratory-based research. The main categories were oncology, neurology, immunology, internal medicine, neurosciences, and public, environmental, and occupational health.

In addition, most of the research done that at the University of Bergen had secured external funding, with the European Union, Research Council of Norway, and the National Institutes of Health in the United States being the leading funders.

## Discussion

Although it is difficult to generalize from our findings to all universities in sub-Saharan Africa, this is to our knowledge one of the first reviews of the newly established universities in this region. Most of the research conducted at the Ethiopian universities (Table 1) focusses on important public health issues, such as children’s health, communicable diseases, and nutritional disorders. Although the production of scientific publications is small, there has been remarkable growth in the number of articles in the past several years. However, the number of publications per author seems to be low, and few researchers demonstrated publication activities over several years.

Although it is laudable that the universities focus on important public health challenges, the lack of research in clinical disciplines is noteworthy. Lack of research training, weak laboratory support, or time constraints due to working in busy clinical departments could explain the dearth of research in clinical disciplines. Furthermore, our review shows that very few studies used randomised controlled trial, cohort, or case-control designs. Thus, most of the research papers were cross-sectional and descriptive studies. Although it is important to describe important health problems relevant for an area, such studies may have limited value in in establishing causal links and thus improving health services for its target populations. Several explanations for these limitations could exist, and the lack of funds to carry out trials or longitudinal studies may have been a limiting factor.

Furthermore, our study shows that only a few research groups functioned over a span of years. These groups were responsible for a large part of the analytical publications, and most of them were funded by external grants. I could also high staff mobility within or out of the country, and institutions may need to strengthen mechanisms to retain their staff.

Unfortunately, our study does not provide sufficient information about the desired collaboration with implementing institutions such as the Ministry of Health, but very few of the co-authors had addresses suggesting that they worked at ministries, hospitals, or public health institutions.

Our bibliometric analysis shows that citations are higher in papers with international collaboration, and the proportion of collaborative research is low as compared to some other Sub-Saharan Africa countries. In some countries the former colonial countries dominate the research agenda.^14^ It should, however, be possible to find a balance in international research collaboration with the aim of enhancing the quality of publications. Others have focused on the obstacles researchers in Africa face in obtaining funds. Many experience that the research funding that reaches institutions in Africa is only a small proportion of the financial support available, and it is important that a larger proportion of the resources is provided directly to African scientists and institutions.^15^ By doing so, this could empower and strengthen their research agenda to conduct research appropriate to the continent.

Furthermore, the research output per author seems to be low, and high turnover of staff, a bureaucratic promotion system, often combined with low salaries, could at least partly explain the low research output. Staff at the universities often work part time for other organisations or in private practices to increase their salaries. Furthermore, many qualified researchers take on administrative responsibilities at their institutions, thereby limiting their time to research. We believe that staff continuity is important, and that staff should be given sufficient time and resources to devote themselves to carrying out research.

Another explanation of the low research output is insufficient national funding for research, and this challenge must be seen in a wider context of the financing of public universities in countries such as Ethiopia.^16^ Furthermore, the universities in our study are young, and may lack an appropriate mixture of junior and senior researchers that are needed to promote the interaction between your researchers and mentors.

## Data Availability

The data for this research are found on PubMed and Web of Science databases. This is publicly available.

https://www.webofscience.com/

## Ethics statement

This study does not involve human participants

## Contributorship statement

### Contributors

BL contributed to the manuscript by making substantial contributions to the conception, acquisition, analysis, and interpretation of data and drafting of the manuscript. FM, TG and SHT contributed to the conception, analysis, and interpretation of data, drafting of the manuscript and approval of the final manuscript. All authors agree to be accountable for all aspects of the work.

## Data availability statement

Data are available upon reasonable request.

## Patient and Public Involvement

No patient involved

